# The acute lag effects of elevated ambient air pollution on stillbirth risk in Ulaanbaatar, Mongolia

**DOI:** 10.1101/2022.02.17.22271117

**Authors:** Temuulen Enebish, David Warburton, Rima Habre, Carrie Breton, Nomindelger Tuvshindorj, Gantuya Tumur, Bayalag Munkhuu, Meredith Franklin

## Abstract

Ulaanbaatar city (UB), the capital and the home to half of Mongolia’s total population, has experienced extreme seasonal air pollution in the past two decades with levels of fine particulate matter with an aerodynamic diameter less than 2.5 micrometers (PM_2.5_) exceeding 500 *μ*g/m^3^ during winter. Based on monitoring data, (PM_2.5_), sulfur dioxide (SO_2_), nitrogen dioxide (NO_2_), and carbon monoxide (CO) exposures were estimated for residential areas across UB using Random Forest models. We collected individual-level data on 1093 stillbirths from UB hospital records (2010-2013) and a surveillance database (2014-2018). Using a time-stratified case-crossover design, we investigated whether short-term increases in daily ambient air pollutants with different exposure lags (2 to 6 days) before delivery were associated with stillbirth. We estimated associations using conditional logistic regression and examined individual-level characteristics for effect modification. During the cold season (Oct-Mar) we observed significantly elevated relative odds of stillbirth per interquartile range increase in mean concentrations of PM_2.5_ (odds ratio [OR]=1.35, 95% confidence interval [CI]=1.07-1.71), SO_2_ (OR=1.71, 95% CI=1.06-2.77), NO_2_ (OR=1.30, 95% CI=0.99-1.72), and CO (OR=1.44, 95% CI=1.17-1.77) 6 days before delivery after adjusting for apparent temperature with a natural cubic spline. The associations of pollutant concentrations with stillbirth were significantly stronger among those younger than 25, nulliparous, and without comorbidities or pregnancy complications during stratified analyses. There was a clear pattern of increased risk for women living in areas of lower socioeconomic status. We conclude that acute exposure to ambient air pollution before delivery may trigger stillbirth, and this risk is higher for certain subsets of women.

## Background

Despite remarkable achievements in women’s and children’s health in the last 15 years, 2.6 million stillbirths in the third trimester occurred worldwide in 2015 (Lawn et al., 2016). Stillbirth was not included in the Millennium Development Goals (United Nations, 2015a) and is still missing from the Sustainable Development goals (United Nations, 2015b). Policies and programs of international and national organizations hardly mention the issue of stillbirth, and it remains an under-financed public health concern. The burden of stillbirth affects women and families by causing psychological and emotional distress and negatively influences communities and society in terms of reduced earnings and increased healthcare expenses. An estimated 4.2 million women live with depression associated with a previous stillbirth (Heazell et al., 2016). Various modifiable risk factors, including maternal age, infectious diseases, lifestyle, and environmental factors, are identified for stillbirth (Lawn et al., 2016). In the past few decades, researchers worldwide have made numerous attempts at ascertaining the effect of environmental exposures such as ambient temperature and air pollution on adverse pregnancy outcomes (Bekkar et al., 2020; Chersich et al., 2020; Klepac et al., 2018). Several reviews summarized the evidence of adverse effects of ambient air pollution on birth outcomes such as preterm birth and low birth weight (Jacobs et al., 2017; Li et al., 2017; Sapkota et al., 2012; Stieb et al., 2012). In contrast, relatively few studies have investigated the association between ambient air pollution and stillbirth. Two recent reviews found suggestive evidence of increased risk of acute and chronic exposure to both gaseous and particulate pollutants on stillbirth occurrence (Siddika et al., 2016; Zhang et al., 2021).

Although the biological mechanism by which ambient air pollutants, notably fine particulate matter with an aerodynamic diameter less than 2.5 micrometers (PM_2.5_), may influence fetal survival is not yet well understood, the literature suggests several processes to describe associations between PM and adverse pregnancy outcomes. One possible pathway indicates that free iron ions on particle surfaces can react with superoxide or hydrogen peroxide to generate highly reactive hydroxyl radicals, which causes excessive oxidative stress that may lead to the degradation of lipids, proteins, and DNA of the placenta (Fortoul et al., 2015). In addition, activation of the inflammatory system may cause PM-mediated oxidative stress. Compounds that adversely affect the immune system promote the release of pro-inflammatory cytokines. In turn, they give a positive feedback loop to form more reactive oxygen species (ROS) and oxidative stress (Al-Gubory et al., 2010). Another potential explanation is that DNA damage induced by pollution may have devastating impacts on a developing fetus whose cells are dividing at a high rate. Studies connecting air pollution to congenital malformations may support this pathway (Padula et al., 2013; Vrijheid et al., 2011). Other studies indicate that air pollution disturbs placental health through injury or inflammation, initiating a severe lack of nutrient transfer between mother and fetus (van den Hooven et al., 2012). To accurately examine the potential mechanistic explanations, additional analyses are required to verify these findings and explore more thoroughly the times during pregnancy when pollutants may impact the fetus resulting in stillbirth.

Ulaanbaatar (UB), the capital city of Mongolia, is in the Tuul river valley between big mountains and resultingly prone to atmospheric inversions in the winter, whereby air pollutants are trapped close to the ground unable to become diluted in the mixing layer of the troposphere. Almost 60% of the city residents used raw coal to heat their home (usually Ger, which is a Mongolian traditional portable dwelling) and cook their meals in cookstoves at the time of this study. This makes UB one of the most polluted cities in the world during winter when there is almost constant atmospheric inversion over the city. Our study area has four major public maternity hospitals that deliver more than 98% of all babies in the city. This situation makes UB a unique location where we can study the acute association between stillbirth and air pollution in a relatively cost-effective and timely manner despite the average stillbirth rate in UB being 7.7 per 1000 total births in 2015 (Lawn et al., 2016).

This paper examined the short-term effect of particulate matter on the risk of stillbirth across lags of 2-6 days. We also investigated the effect modification of these associations by socioeconomic risk factors of stillbirth.

## Methods

Ulaanbaatar city registered about 36 000 deliveries and 250 stillbirths on average per year between 2010 and 2018 across nine districts, which consist of a varying number of Khoroos (minor administrative units of the city). The current study includes stillbirths occurring between January 1, 2010, and December 31, 2018, in 6 contiguous districts (138 Khoroos) of UB. The Institutional Review Boards at the Health Sciences Campus of USC, the Children’s Hospital Los Angeles, and the Medical Ethics Committee of the Ministry of Health of Mongolia approved the study design and methods.

### Study population

We used two different data sources to collect individual-level stillbirth data. Between 2010 and 2013, records were only available at maternity hospital archives in the form of a birth record as there was no official surveillance of stillbirth during this period. In order to collect these data, we hired and trained eight research assistants to help with data abstraction from the paper-based birth records into a computer database. In August 2017, we abstracted the stillbirth data using four two-person teams where one person read paper records, and the other person entered them into the database. Each team member double-checked every record entry. The National Center for Maternal and Child Health (NCMCH) of Mongolia established a Department of Surveillance that conducts nationwide surveillance on maternal and child health indicators in 2014. They use specific forms for each maternal and child health endpoint to collect data. We obtained individual-level stillbirth data between 2014 and 2018 using the department’s electronic database of stillbirth forms. See Supplemental Materials for the translated version of the form.

We abstracted the following information from the birth records and the stillbirth form: maternal age, residential address, occupation, gravidity, parity, prenatal care status, delivery hospital, date of stillbirth, fetal gender, weight, length, and gestational age. We used the best obstetric estimate variable on the birth record to determine gestational age, which combines the last menstrual period and ultrasound parameters, as is commonly accepted in clinical practice for gestational age estimation. The Ministry of Health of Mongolia uses the following definition for stillbirth: the birth of a fetus with ≥500 grams weight at or beyond the 22nd week of gestation and who had no signs of life such as heartbeat, umbilical cord pulse, or muscle movement at birth (Ministry of Health, Guideline for the Calculation and Definition of Main Health Indicators, 2004, page 19). We included stillbirths based on the following criteria: singleton status, maternal residence in one of the six central districts, and delivery at four major public maternity hospitals (Figure 1). We excluded intrapartum stillbirths based on biological plausibility: the definition being a fetal death occurring after the onset of labor and prior to delivery (Tavares Da Silva et al., 2016). Based on the residential Khoroo of each subject, we also obtained Khoroo-level variables such as percentage of households with internet connectivity (a proxy for socioeconomic status) and percentage of Ger households from the UB Office of Statistics, and the number of stoves per square kilometer from the UB Office of Air Pollution Reduction (Narmandakh et al., 2018).

**Figure 1.**
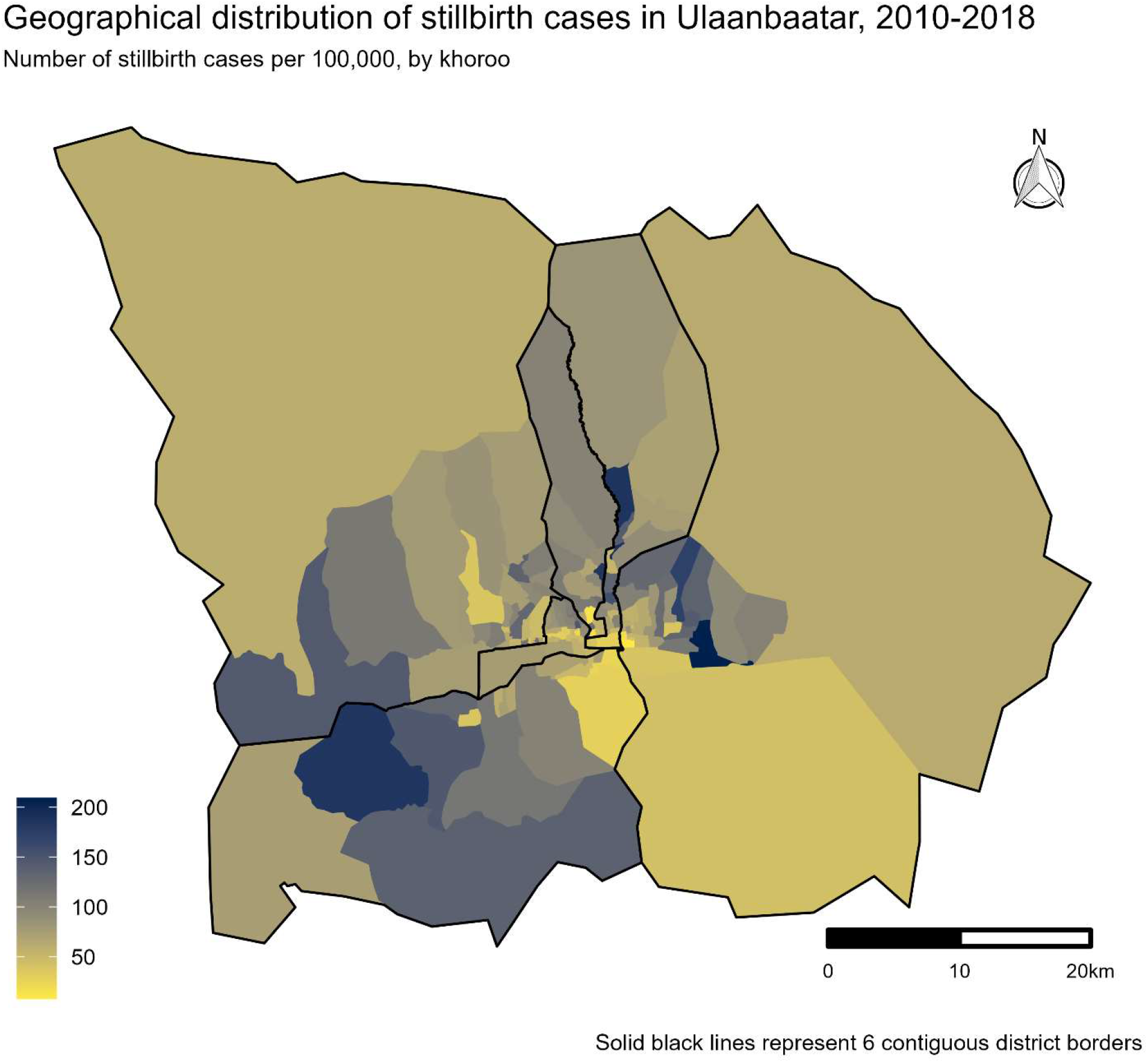
Ulaanbaatar city map of 6 main contiguous districts, the colors represent the rate of stillbirth cases

### Exposure data

We linked daily air pollution concentrations that were estimated using a previously developed and validated spatiotemporal model (Enebish et al., 2020) to the residential Khoroos and for the relevant dates of each stillbirth case. Briefly, our model used ground-level measurements from stationary stations supplemented with multiple spatiotemporal covariates including meteorological (atmospheric temperature, relative humidity, wind speed, wind direction), land use (length of roads) as well as population (Ger households, stove density), and indicators for time (day of year, season, month). We evaluated six different machine learning algorithms and found that decision trees, particularly Random Forest models, performed the best with leave-one-location-out cross-validation R^2^ of 0.82 and hold-out test R^2^ of 0.96 for PM_2.5_. These Random Forest models were used to predict daily PM_2.5_, SO_2_, NO_2_, and CO concentrations at the centroid of each Khoroo of UB city between 2010 and 2018. In addition, we retrieved meteorological variables from two stations (Chinggis Khaan Intl, Songiin) in UB that send their data to the Global Hourly Integrated Surface Dataset of the National Oceanic and Atmospheric Administration (NOAA National Centers for Environmental Information, n.d.). Using these data, we calculated Apparent Temperature (AT) as either a wind chill index or a heat index depending on the temperature. Wind chill combines temperature and wind velocity to capture how cold the weather feels to the average person when the temperate falls below 0°C. We used equations used by Environment and Climate Change Canada (Nelson et al., 2002) since Mongolian National Agency for Meteorology and Environmental Monitoring (NAMEM) currently does not have a formula adopted. Depending on air temperature and wind speed, we applied Equation 1 when the temperature of the air was ≤ 0°C and the reported wind speed was ≥ 5 km/h, and Equation 2 when the temperature of the air was ≤ 0°C and the reported wind speed was > 0 km/h but < 5 km/h.

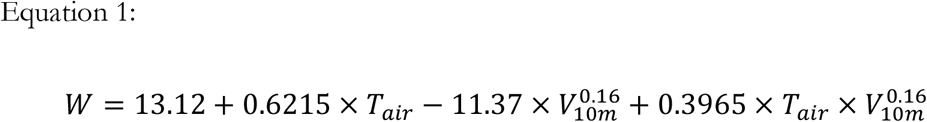

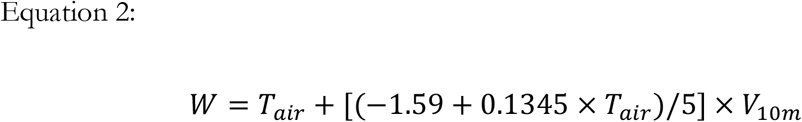

Where *W* is the wind chill index in degrees Celsius, *T*_air_ is the air temperature in degrees Celsius (°C), and *V*_10m_ is the wind speed at 10 meters (standard anemometer height), in kilometers per hour (km/h).

The heat index measures heat exposure and combines temperature and relative humidity to obtain perceived temperature when the weather is hot. We used the US National Weather Service algorithm implemented by the R package “weathermetrics” (Anderson et al., 2013) to calculate AT when the atmospheric temperature was more than 5°C.

### Study design

Using a time-stratified case-crossover design, we estimated the relative odds of stillbirth associated with each interquartile range (IQR) increase in mean pollutant concentrations. Maclure first proposed the case-crossover design to study the transient effect of exposure on rare-acute onset disease (Maclure, 1991). Conceptually, the design resembles a combination of retrospective, nonrandomized crossover design and matched case-control design. The main difference is that the case-crossover design only uses a sample of the base population-time by including only cases to allow control of time-independent confounders within subjects. The design derives the effect estimates by comparing exposure before the event with exposure at other control (referent) times. Choosing referent times in case-crossover studies of air pollution is particularly important due to time-dependent confounders, time trends, and autocorrelation in the air pollution exposure. The time-stratified referent scheme is the only referent selection that can avoid overlap bias due to conditional logistic regression estimating equations and time-trend bias by selecting random index dates based on stratification (Janes et al., 2005). We stratified on year, month, and day of the week to control long-term, seasonal, and daily trends using time-stratified referent selection. We selected referent (control) periods as every seventh day during the same month as the stillbirth (case period). For instance, if the case period was on Wednesday, every other Wednesday in the same month was considered a control period. We looked at lag days between 2 and 6 as possible case periods because it may take an average of 48-70 hours for the deceased fetus to be expelled from the womb (Gardosi et al., 1998; Genest et al., 1992).

### Statistical analysis

We utilized conditional logistic regression to estimate the odds ratios (OR) of delivering stillbirth for every IQR increase in PM_2.5_ on lag days 2 to 6. Models were estimated using a stratified Cox model, with each subject assigned to its stratum (Gail et al., 1981). We adjusted each model with a natural cubic spline (4 degrees of freedom) of the mean apparent temperature on corresponding lag days to control for time-variant meteorological factors and their non-linear relationship with stillbirth as in previous studies (Faiz et al., 2013). We also conducted a seasonally stratified analysis, restricting the models to cases that occurred only in the cold season (October-March), considering the disproportional seasonal variation caused by air pollution emission sources in UB during cold weather (Figure 2). The same models were fit to SO_2_, NO_2_, and CO with their ORs and 95% confidence intervals (CIs) scaled to the IQR of each pollutant. We ran stratified analyses based on the following variables to evaluate differential risks of stillbirth by individual case characteristics: maternal age, employment, number of previous pregnancies and deliveries, comorbidity, pregnancy complication as well as residential level covariates such as percentage of Ger households and households with internet connectivity in each subject’s living Khoroo. All statistical modeling and geospatial computing were performed in R version 3.6.2 (R Core Team, 2019) using packages “tidyverse” (Wickham et al., 2019), “sf” (Pebesma, 2018), and “survival” (Therneau & Grambsch, 2000).

**Figure 2.**
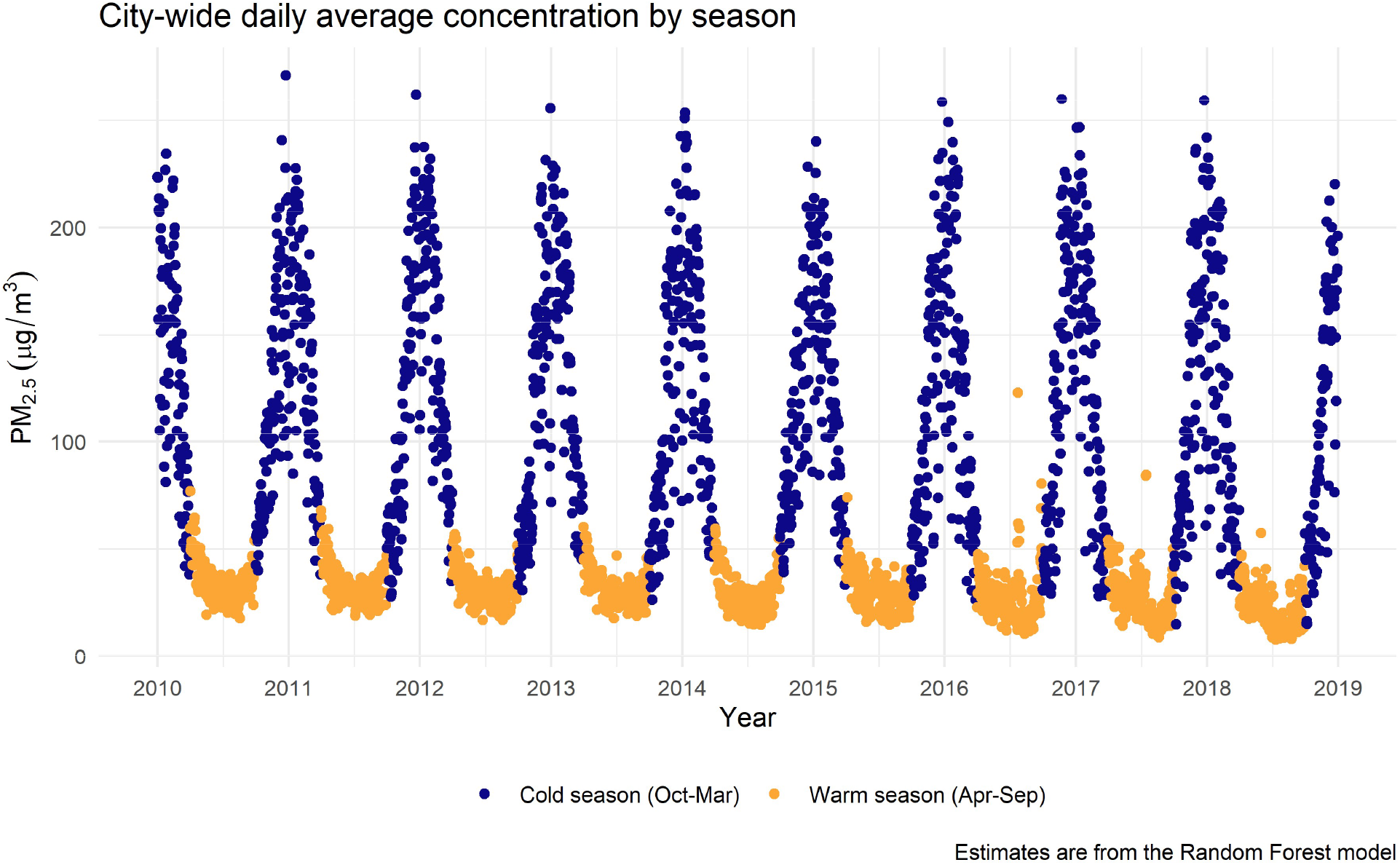
Seasonal variation of modeled PM_2.5_ concentration in UB

## Results

After applying inclusion and exclusion criteria, our study population consisted of 1093 stillbirth cases between 2010 and 2018 (Figure 3).

**Figure 3.**
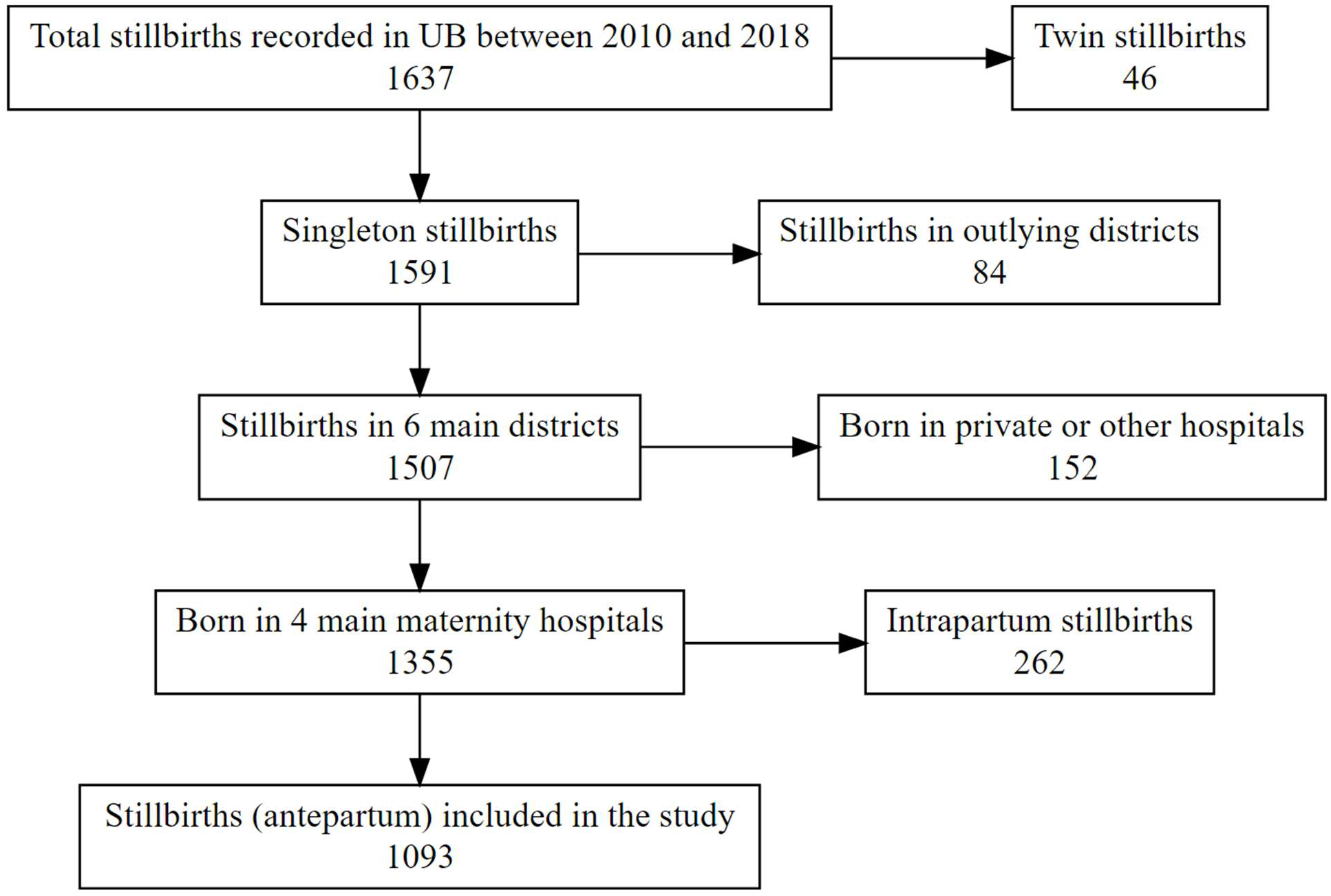
Flow chart of the study population

Almost a quarter of stillbirths occurred among women aged 35 years or older, and the subjects were divided equally among employed and unemployed status. More than half (54%) of the women had been pregnant more than two times; however, a third (35%) of the women had never given birth before. About 65% of the subjects had one or more pregnancy complications, and approximately half of them had various comorbidities. In terms of fetal variables, there was no dominant fetal gender. The majority weighed less than 2500 grams and had gestational weeks less than 36 weeks (Table 1). Table 1 shows that majority of the cases came from Khoroos with lower internet connectivity, more Ger households, and higher stove density.

**Table 1.**
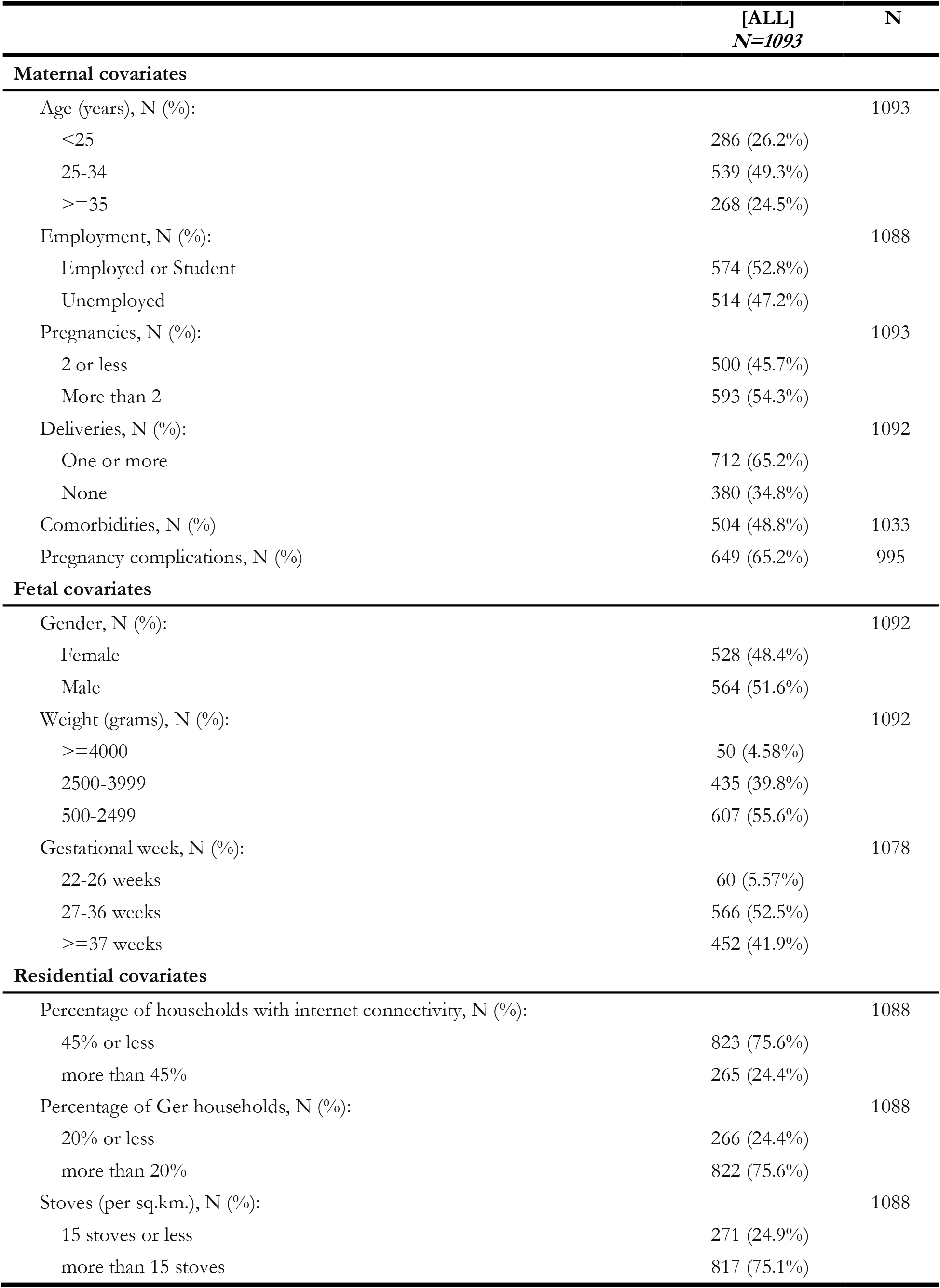
Characteristics of Mothers and Stillbirths, Ulaanbaatar 2010–2018

|  | [ALL]<br>N=1093 | N |
| --- | --- | --- |
| **Maternal covariates** |  |  |
| Age (years), N (%): |  | 1093 |
| <25 | 286 (26.2%) |  |
| 25-34 | 539 (49.3%) |  |
| ≥35 | 268 (24.5%) |  |
| Employment, N (%): |  | 1088 |
| Employed or Student | 574 (52.8%) |  |
| Unemployed | 514 (47.2%) |  |
| Pregnancies, N (%): |  | 1093 |
| 2 or less | 500 (45.7%) |  |
| More than 2 | 593 (54.3%) |  |
| Deliveries, N (%): |  | 1092 |
| One or more | 712 (65.2%) |  |
| None | 380 (34.8%) |  |
| Comorbidities, N (%) | 504 (48.8%) | 1033 |
| Pregnancy complications, N (%) | 649 (65.2%) | 995 |
| **Fetal covariates** |  |  |
| Gender, N (%): |  | 1092 |
| Female | 528 (48.4%) |  |
| Male | 564 (51.6%) |  |
| Weight (grams), N (%): |  | 1092 |
| ≥4000 | 50 (4.58%) |  |
| 2500-3999 | 435 (39.8%) |  |
| 500-2499 | 607 (55.6%) |  |
| Gestational week, N (%): |  | 1078 |
| 22-26 weeks | 60 (5.57%) |  |
| 27-36 weeks | 566 (52.5%) |  |
| ≥37 weeks | 452 (41.9%) |  |
| **Residential covariates** |  |  |
| Percentage of households with internet connectivity, N (%): |  | 1088 |
| 45% or less | 823 (75.6%) |  |
| more than 45% | 265 (24.4%) |  |
| Percentage of Ger households, N (%): |  | 1088 |
| 20% or less | 266 (24.4%) |  |
| more than 20% | 822 (75.6%) |  |
| Stoves (per sq.km.), N (%): |  | 1088 |
| 15 stoves or less | 271 (24.9%) |  |
| more than 15 stoves | 817 (75.1%) |  |

The models looking at the overall period showed a consistently increased risk of stillbirth on 3 and 6 days before delivery for all pollutants examined (Table 2 and Figure 4). Odds ratios on lag day 6 were generally higher than the estimates on lag day 3. Only carbon monoxide had a statistically significant elevated risk on lag day six at *α*=0.05 significance level. In models restricted to the cold season, most effect estimates except NO_2_ increased considerably. Previously non-significant odds ratios on lag day 6 in overall models reached statistical significance for PM_2.5_ (OR:1.35, 95% CI:1.07-1.71) and SO_2_ (OR:1.71, 95% CI:1.06-2.77). We also found a statistically significant increased risk of stillbirth per IQR increase in mean PM_2.5_ 3 days before delivery (Figure 4).

**Table 2.** Relative Odds of Stillbirth Associated with IQR Increases in Mean PM_2.5_, NO_2_, SO_2_, and CO Concentration by Lag day(s)

| Pollutants | Lag Days | IQR, µg/m<sup>3</sup> | OR<sup>1</sup> | (95% CI)<sup>2</sup> | OR | (95% CI) |
| --- | --- | --- | --- | --- | --- | --- |
|  |  |  | Overall (n = 1 088)<sup>3</sup> |  | Cold season (n = 515)<sup>3</sup> |  |
| PM<sub>2.5</sub> | 2 | 74.67 | 0.99 | (0.8-1.23) | 0.96 | (0.76-1.2) |
|  | 3 | 77.00 | 1.23 | (0.98-1.54) | 1.28 | (1-1.62) |
|  | 4 | 75.24 | 0.92 | (0.74-1.14) | 0.95 | (0.76-1.19) |
|  | 5 | 74.65 | 1.01 | (0.81-1.26) | 1.05 | (0.83-1.31) |
|  | 6 | 75.72 | 1.24 | (0.99-1.56) | 1.35 | (1.07-1.71) |
| SO<sub>2</sub> | 2 | 36.40 | 0.94 | (0.61-1.44) | 0.84 | (0.53-1.33) |
|  | 3 | 37.12 | 1.26 | (0.81-1.97) | 1.33 | (0.82-2.14) |
|  | 4 | 35.82 | 0.94 | (0.61-1.46) | 1.02 | (0.64-1.63) |
|  | 5 | 35.73 | 0.87 | (0.56-1.36) | 1.00 | (0.63-1.61) |
|  | 6 | 36.32 | 1.47 | (0.95-2.3) | 1.71 | (1.06-2.77) |
| NO<sub>2</sub> | 2 | 29.89 | 1.01 | (0.79-1.29) | 0.89 | (0.68-1.16) |
|  | 3 | 30.55 | 1.18 | (0.92-1.52) | 1.14 | (0.86-1.5) |
|  | 4 | 29.73 | 1.02 | (0.8-1.29) | 0.94 | (0.72-1.23) |
|  | 5 | 30.12 | 1.14 | (0.89-1.45) | 1.12 | (0.85-1.47) |
|  | 6 | 30.14 | 1.21 | (0.95-1.55) | 1.30 | (0.99-1.72) |
| CO | 2 | 561.03 | 1.03 | (0.87-1.24) | 1.05 | (0.86-1.27) |
|  | 3 | 572.20 | 1.18 | (0.98-1.42) | 1.21 | (0.99-1.47) |
|  | 4 | 560.32 | 1.01 | (0.85-1.21) | 1.04 | (0.86-1.26) |
|  | 5 | 556.54 | 1.00 | (0.83-1.19) | 1.04 | (0.86-1.27) |
|  | 6 | 571.54 | 1.25 | (1.04-1.51) | 1.44 | (1.17-1.77) |

<sup>1</sup>Odds Ratio
<sup>2</sup>95% Confidence Interval
<sup>3</sup>Models are adjusted for the apparent temperature of the corresponding lag day

**Figure 4.**
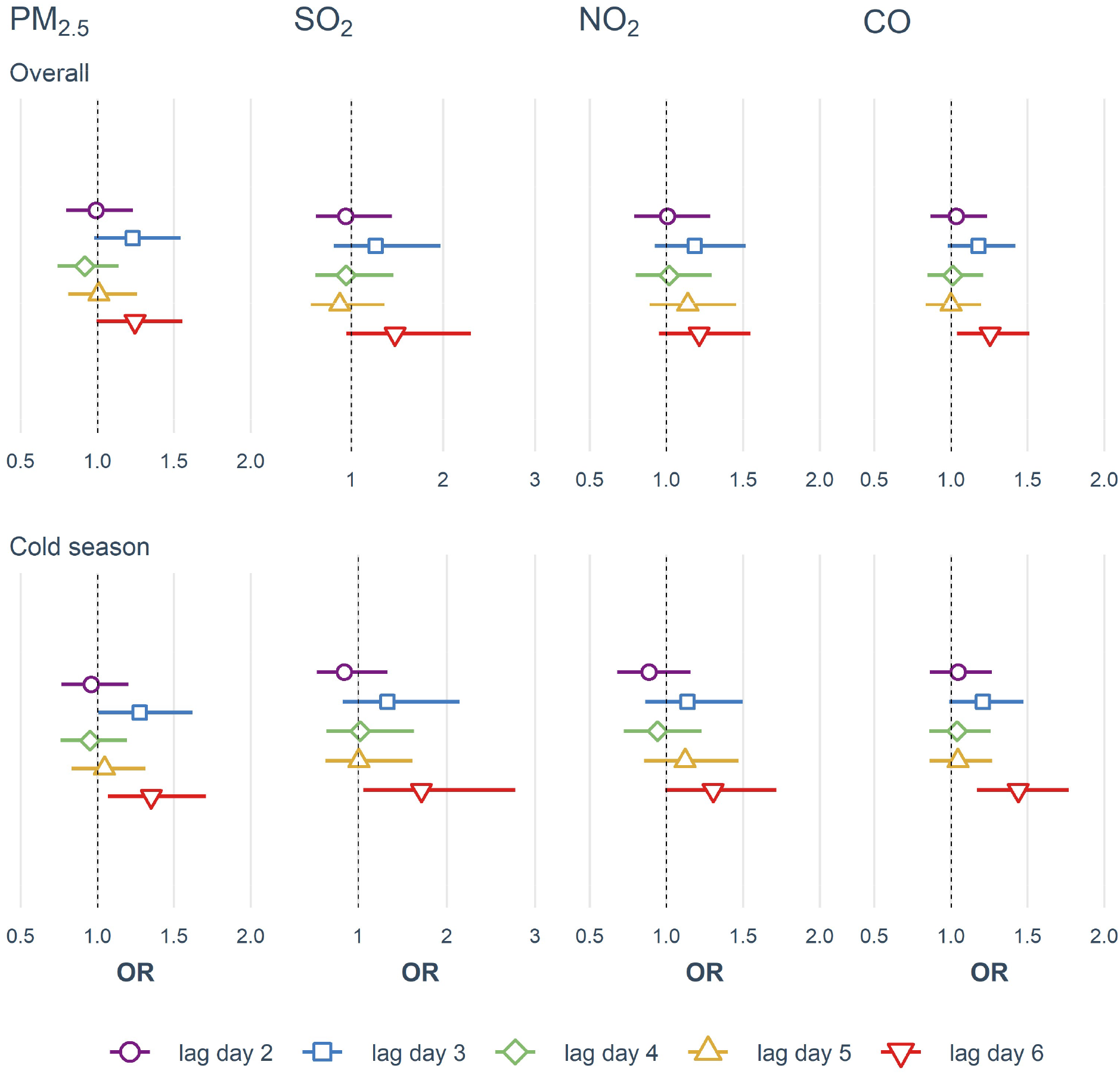
Effect estimates of air pollution on stillbirth risk

Possible modifiers of the effect associated with stillbirth and pollutants examined in stratified models (Table 3) show significant differences in odds ratios between various maternal and residential characteristics. They reveal stronger associations for younger, unemployed women with fewer pregnancies and women who had never given birth before. Interestingly, we also found higher odds ratios for women without comorbidities and pregnancy complications. On the other hand, living in a Khoroo with a larger proportion of Ger households and households without internet connectivity and higher stove density was associated with higher relative odds of stillbirth.

**Table 3.** Relative Odds of Stillbirth Associated with IQR Increase in PM_2.5_, NO_2_, SO_2_, and CO Concentration on Lag Day 6 by Level of Maternal Characteristics

|  | PM<sub>2.5</sub> |  | SO<sub>2</sub> |  | NO<sub>2</sub> |  | CO |  |
| --- | --- | --- | --- | --- | --- | --- | --- | --- |
| Characteristic | OR | (95% CI) | OR | (95% CI) | OR | (95% CI) | OR | (95% CI) |
| **Maternal age (years)** |  |  |  |  |  |  |  |  |
| <25 | 1.45 | (0.94-2.25) | **2.38** | **(1.02-5.57)** | 1.22 | (0.8-1.88) | **1.52** | **(1.04-2.21)** |
| 25-34 | 1.19 | (0.86-1.66) | 1.25 | (0.65-2.4) | 1.20 | (0.83-1.73) | 1.21 | (0.91-1.6) |
| ≥35 | 1.17 | (0.77-1.78) | 1.20 | (0.51-2.83) | 1.21 | (0.73-2.02) | 1.15 | (0.81-1.62) |
| **Maternal employment** |  |  |  |  |  |  |  |  |
| Unemployed | **1.43** | **(1.01-2.03)** | 1.89 | (0.98-3.62) | **1.53** | **(1.06-2.2)** | **1.49** | **(1.11-1.98)** |
| Employed or Student | 1.12 | (0.83-1.51) | 1.19 | (0.64-2.2) | 0.98 | (0.7-1.37) | 1.12 | (0.87-1.43) |
| **Number of pregnancies** |  |  |  |  |  |  |  |  |
| 2 or less | **1.67** | **(1.19-2.34)** | **2.59** | **(1.33-5.03)** | **1.40** | **(1-1.96)** | **1.54** | **(1.15-2.04)** |
| More than 2 | 0.98 | (0.73-1.33) | 0.92 | (0.51-1.68) | 1.04 | (0.73-1.48) | 1.07 | (0.84-1.38) |
| **Number of deliveries** |  |  |  |  |  |  |  |  |
| None | **1.74** | **(1.2-2.52)** | **2.76** | **(1.32-5.77)** | 1.39 | (0.96-2.01) | **1.64** | **(1.2-2.23)** |
| 1 or more | 1.02 | (0.77-1.36) | 1.00 | (0.57-1.76) | 1.08 | (0.78-1.49) | 1.07 | (0.84-1.35) |
| **Maternal comorbidity** |  |  |  |  |  |  |  |  |
| Yes | 1.13 | (0.83-1.55) | 1.07 | (0.56-2.05) | 1.13 | (0.81-1.6) | 1.25 | (0.96-1.62) |
| No | **1.39** | **(1-1.93)** | **1.97** | **(1.05-3.7)** | 1.23 | (0.86-1.77) | 1.25 | (0.95-1.64) |
| **Pregnancy complication** |  |  |  |  |  |  |  |  |
| Yes | 1.12 | (0.84-1.5) | 1.17 | (0.67-2.03) | 1.07 | (0.79-1.46) | 1.05 | (0.82-1.34) |
| No | 1.46 | (0.98-2.19) | **2.44** | **(1.07-5.58)** | **1.56** | **(1-2.44)** | **1.57** | **(1.11-2.23)** |
| **Percentage of households with internet connection** |  |  |  |  |  |  |  |  |
| 45% or less | **1.37** | **(1.05-1.78)** | **1.81** | **(1.1-2.97)** | 1.29 | (0.98-1.69) | **1.41** | **(1.13-1.77)** |
| more than 45% | 0.99 | (0.65-1.51) | 0.53 | (0.19-1.5) | 0.86 | (0.49-1.53) | 0.94 | (0.65-1.35) |
| **Percentage of Ger households** |  |  |  |  |  |  |  |  |
| 20% or less | 1.10 | (0.72-1.68) | 0.80 | (0.28-2.28) | 1.04 | (0.62-1.74) | 0.89 | (0.61-1.3) |
| more than 20% | **1.32** | **(1.01-1.72)** | **1.63** | **(1-2.68)** | 1.25 | (0.94-1.65) | **1.42** | **(1.14-1.77)** |
| **Stoves (per sq.km.)** |  |  |  |  |  |  |  |  |
| 15 stoves or less | 0.93 | (0.61-1.44) | 0.58 | (0.2-1.66) | 0.81 | (0.49-1.35) | 0.83 | (0.56-1.23) |
| more than 15 stoves | **1.39** | **(1.07-1.81)** | **1.77** | **(1.08-2.9)** | **1.37** | **(1.03-1.82)** | **1.42** | **(1.14-1.77)** |

Estimates significant at the 0.05 significance level are **bolded**

## Discussion

We found a substantially increased risk of stillbirth associated with IQR increases in the mean concentrations of all examined pollutants 3 and 6 days before delivery. These associations were controlled for time-varying confounders such as atmospheric temperature, relative humidity, and wind velocity in the form of apparent temperature. Under a time-stratified case-crossover design, time-invariant confounders such as individual-level maternal, fetal, and residential attributes and seasonal, monthly, and day-of-the-week time trends could not have influenced our results. Our stratified analyses showed a disproportionately higher risk for women living in low-income Ger districts than in apartments. They also indicate that younger women who have not given birth before and have no comorbidities and pregnancy complications are at increased risk of stillbirth associated with ambient air pollution.

The biological mechanisms underlying the effect of air pollutants on stillbirth risk have varying levels of evidence depending on the pollutant. On the one hand, there is well-established toxicity of carbon monoxide on the fetus (Penney, 1996). Two main pathways are reduction of oxygen-carrying capacity of maternal hemoglobin by CO, which leads to oxygen shortage in fetal blood (Salam et al., 2005), and higher affinity of fetal hemoglobin for binding CO than adult hemoglobin that further compromises the oxygen delivery (Sangalli et al., 2003). On the other, we have scarce evidence of how exactly NO_2_ and SO_2_ might affect fetal death. There is evidence for crossing the placental barrier, affecting cell division, and activating hypoxic injury or immune-mediated damage (Al-Gubory et al., 2010; Proietti et al., 2013). There have been several hypotheses regarding their effect on a fetus for particulate matter. Major ones are a compromise of maternal blood and nutrients delivery, DNA damage and inflammation via contribution to oxidative stress, and lowering the transplacental function by increasing the concentration of DNA adducts (Perera et al., 1992). Timing of exposure to particulate matter along fetal developmental periods may also lead to varying effects due to differences in physiologic maturity of the fetus (Perera et al., 1999).

While numerous studies have examined the association between air pollutants and adverse pregnancy outcomes, relatively few investigated the effect on stillbirth. Even fewer studies focused on the acute effects of air pollution on stillbirth risk. A time-series study in Sao Paulo, Brazil, examined the association between daily counts of intrauterine mortality (gestational duration of >28 weeks) and ecological measures of daily ambient air pollutants. They found increased short-term (less than five days) risk of intrauterine mortality for NO_2_ (*β*=0.0013/*μ*g/m^3^; *p*<0.01), SO_2_ (*β*=0.0005/*μ*g/m^3^; *p*<0.10) and CO (*β*=0.0223/ppm; *p*<0.10) using Poisson regression adjusted for season and weather (Pereira et al., 1998). This study, to our knowledge, is the first to look at the short-term effect of air pollutants on stillbirth risk and remains an essential part of the evidence base despite its limitations due to ecological design and imperfect case definition.

A study by Faiz et al. utilized time-stratified referent selection in a case-crossover design to look at the triggering effect of ambient air pollution on stillbirth in New Jersey, US (Faiz et al., 2013). They found significantly increased relative odds of stillbirth on lag day 2 per IQR increase in mean concentrations of CO (OR = 1.20, 95% CI = 1.05-1.37) and SO_2_ (OR = 1.11, 95% CI = 1.02-1.22) and increased odds ratios for IQR increases in NO_2_ (OR = 1.11, 95% CI = 0.97–1.26) and PM_2.5_ (OR = 1.07, 95% CI = 0.93–1.22) levels. Similar increases in stillbirth risk were also observed for cumulative averages on days 2 to 6, and they did not observe effect modifications by maternal characteristics. The current study tries to emulate the above study conceptually to investigate the acute effect of ambient air pollutants on the risk of stillbirth. Our findings agree on increased short-term risk of stillbirth associated with IQR increase in the pollutants. They, however, differ in terms of which lag day has the highest risk and the magnitude of the risk. We found increased relative odds of stillbirth on lag days 3 and 6 for all pollutants, and the ORs were amplified when restricted to the cold season only. Unfortunately, there was almost no overlap between individual maternal risk factors between the studies to compare stratified analyses.

Three recent studies on the short-term association between stillbirth and ambient pollutants have been published. Mendola et al. used retrospective cohort data across 12 clinical centers in the US. They investigated the acute and chronic effects of air pollutants on the risk of stillbirth at the community level using Poisson regression with generalized estimating equations. They observed that acute exposure to ozone was associated with a 13-22% increased risk of stillbirth on lag days 2, 3, and 5-7 (Mendola et al., 2017). A time-series study in Ahvaz, Iran, by Dastoorpoor et al. used a distributed lag non-linear model estimated by quasi-Poisson regression to investigate the acute effects of air pollution (per 10-unit increase) on stillbirth and other adverse pregnancy outcomes.

They looked at single-day lags of 1 and 2 and cumulative lag days 0 through 14 and did not find an increased effect estimate, but rather an inverse association with ozone on lag day 1 and 2 and with SO_2_ on lag day 2 (Dastoorpoor et al., 2018). A recent addition to the evidence base on the short-term effects of ambient air pollution on stillbirth risk came from California. They used population-weighted centroids to link ground monitoring station data to the maternal residential zip code to assign air pollution exposure. Similar to New Jersey and our study, the authors utilized a time-stratified case-crossover design to estimate the short-term effects and found an increased risk of stillbirth per IQR increase in SO_2_, O_3_, and PM_10-2.5_ concentrations (Sarovar et al., 2020).

Our findings are broadly consistent with previous literature on the topic; however, we must acknowledge several differences. The magnitude of the effect that we observed is substantially larger than the studies mentioned above. We believe this is primarily because of the large IQR stemming from high air pollution levels during cold months in UB. Unfortunately, this change in daily air quality over days and weeks during winter in UB is in line with the significant health and mortality burden of air pollution found in previous studies. Allen et al. estimated conservatively that 1 in 10 deaths in UB are due to air pollution using land-use regression models and mobile monitoring to assess exposure (Allen et al., 2013). At the same time, researchers from the National Center for Maternal and Child Health (NCMCH) found a strikingly high seasonal correlation between miscarriage and air pollution levels in the city (Enkhmaa et al., 2014).

The main improvement of our study over previous literature comes from our individual-level exposure assessment with fine spatial and temporal resolution. The main challenge in conducting air pollution epidemiology studies in low to middle income countries is the difficulty of accurately assessing exposure due to inadequate and unreliable measurements provided by scarce pollutant monitoring stations. Our previous work demonstrated the feasibility and advantages of utilizing machine learning algorithms to capture complex relationships between air pollution and other meteorological, land use, and population-level variables in a low-resource setting with sparse monitoring capacity (Enebish et al., 2020). This work allowed us to assign air pollution exposure to every subject in our study based on their residential area and relevant days. Although exposure measurement errors are unavoidable when we try to estimate personal exposure using modeling based on ambient air pollution levels, we believe this approach has significantly improved over previous methods of assigning exposures. But the error will result in non-differential misclassification, likely leading to a bias towards the null. We should also mention that we evaluated various modifiable maternal risk factors that previous studies have not been able to investigate. This opportunity is mainly thanks to the manual data collection process from birth records and stillbirth form information provided by the Surveillance Department of NCMCH. The limitations of our current study include exposure assessment at the residential area (Khoroo) level, the uncertain estimate for the date of fetal death based on the delivery date, and possible residual confounding due to imperfect measurement of apparent temperature.

In conclusion, we found a considerably increased risk of stillbirth on days 3 and 6 before delivery for every IQR increase in mean concentrations of PM_2.5_, SO_2_, NO_2_, and CO. This association was strengthened when we restricted the analysis to only the cold season. Future studies with finer spatial and temporal exposure, time-varying confounder assessment, and better case identification are necessary to elucidate further the underlying mechanism of the markedly adverse effects of air pollution on stillbirth.

## Data Availability

All data produced in the present study are available upon reasonable request to the authors.

